# Understanding Knowledge, Attitudes and Practices on Ebola Virus Disease: A Multi-Site Mixed Methods Survey on Preparedness in Rwanda

**DOI:** 10.1101/2022.11.03.22281885

**Authors:** Janvier Karuhije, Menelas Nkeshimana, Fathiah Zakham, Benjamin Hewins, Justin Rutayisire, Gustavo S. Martinez, David Kelvin, Pacifique Ndishimye

**Affiliations:** Rwanda Biomedical Centre, Ministry of Health, Kigali, Rwanda; University Teaching Hospital of Kigali, Kigali, Rwanda; Department of Virology, Faculty of Medicine, University of Helsinki, Helsinki, Finland; Laboratory of Emerging Infectious Diseases, Department of Microbiology and Immunology, Canadian Centre for Vaccinology CCfV, Faculty of Medicine, Dalhousie University, Halifax, Canada; United Nations Children’s Fund (UNICEF), Kigali, Rwanda

## Abstract

The overall goal of this survey was to understand the Ebola Virus Disease (EVD) - related knowledge, attitudes, and practices (KAP) at individual, inter-personal, institutional, and societal levels in Rwanda. This cross-sectional mixed-methods survey was conducted in five selected districts: Rusizi, Karongi, Rubavu, Burera and Gasabo. Quantitative data was collected from 1,010 participants using a structured questionnaire and Kobo Collect. Qualitative data was collected from 98 participants through Key Informant Interviews and Focus Group Discussions using a semi structured interview guide. Among the 1,010 surveyed respondents, 56% were male, 70.3% were married, and 50% had primary education. An important finding was the high level of Ebola awareness and knowledge in all the five districts, with 99.6% reporting having previously heard of Ebola, which indicates previous awareness-raising efforts were successful. More than 54% of respondents indicated that Ebola is caused by a virus which originates from wild animal animals (42.1%). Furthermore, fever (85%), bleeding (87.7), and vomiting (40.2%) were cited as the primary signs and symptoms for Ebola. Most of the respondents were knowledgeable regarding prevention measures for Ebola. Despite this, 80% of the survey respondents had not received formal training or health education on Ebola. The majority of respondents (78.2%) reported having a positive attitude towards EVD survivors. Many respondents (90%) believe that the country is at risk of an EVD outbreak and about 87.8% think that they are personally at risk of contracting Ebola. Most respondents reported adopting habits that included avoiding physical contact with the patients and reducing unnecessary movements/travel throughout the Ebola-affected regions. At the community level, participants state that they participate in the sharing of Ebola-related information and reporting suspected cases to relevant authorities. Additionally, many participants know the necessary emergency contact number (114) for assistance and reporting of EVD-related information. Most respondents (97.2%) believed that it is important to be vaccinated to prevent Ebola, and around 93.3% are ready/willing to be vaccinated once the EVD vaccine is available. While the radio is the preferred source for Ebola-related information, the most trusted sources are the ministry of health and governmental institutions, such as the Rwanda Biomedical Centre. Our results show that there was high EVD-related knowledge and awareness among the general population in Rwanda. However, for strong public health awareness, preparedness, and protection, there is a need to implement public sensitization programmes that address EVD-related misconceptions and discriminatory attitudes toward EVD patients.

## Introduction

Ebola Virus disease (EVD), formerly known as Ebola hemorrhagic fever, is a severe and often fatal illness that affects humans. It was first described in 1976 near the Ebola River in what is now the Democratic Republic of the Congo(Centers for Disease Control and Prevention, 2020). The average EVD case fatality rate is around 50% and the case fatality rates have varied from 40% to 90% in past outbreaks (Gupta et al., 2021; Hamid et al., 2018; World Health Organization, 2021b). Originally transmitted to humans from wild animals, the spread of EVD is driven by human-to-human transmissions (WHO, 2018). Consequently, community engagement is pivotal in the fight against EVD and successfully containing and controlling outbreaks. For effective outbreak control, the provision of comprehensive interventions including case management; infection prevention and control practices; surveillance and contact tracing; reliable laboratory services; safe and dignified burials and social mobilization are required (Gupta et al., 2021; Olu et al., 2016). According to the World Health Organization (WHO) (2020), EVD vaccines have been used to help control the spread of Ebola outbreaks in Guinea and in the Democratic Republic of Congo (DRC). There are currently two treatments approved by the U.S. Food and Drug Administration (FDA) to treat EVD caused by the Ebola virus, species *Zaire ebolavirus*, in adults and children. The first drug approved in October 2020, Inmazeb™, is a ‘cocktail’ combination of three monoclonal antibodies. The second drug, Ebanga™, is a single monoclonal antibody and was approved in December 2020 (CDC, 2019; World Health Organization, 2022a).

The 2014 (to 2016) EVD outbreak that affected Guinea, Liberia and Sierra Leone was the longest, deadliest, and most complex outbreak since the discovery of the virus in 1976 (Olu et al., 2016; Shoman et al., 2017). The outbreak has not yet fully abated, and to date the death-toll stands at 11,000 people at an estimated total cost of US$4.3 billion. Guinea subsequently declared a new Ebola outbreak in February 2021, mandating the establishment of preventative measures among its neighboring countries (World Health Organization, 2021a).

Since August 2018, the Democratic Republic of the Congo (DRC) has been facing a large-scale EVD outbreak in the eastern provinces of North Kivu and Ituri, with the outbreak also spreading to Goma and South Kivu provinces. This EVD outbreak is the country’s 10^th^ in recorded history and has infected 3,303 people and resulted in 2,199 deaths from August 2018 to November 2019, according to data from the WHO. By November 2019, 28% of the cases recorded during the outbreak were reported in children, representing the largest child EVD burden in recorded history. The unprecedented growth in cases (particularly in children) prompted WHO to declare (July 2019) the Ebola outbreak in DRC a Public Health Emergency of International Concern (Eurosurveillance Editorial Team, 2019; WHO, 2019). From 1 June 2020 to 18 November 2020, a total of 130 EVD cases including 119 confirmed and 11 probable cases were reported in the country. In February 2021, a new EVD case was detected in Butembo, a city in DRC’s North Kivu Province. Unstable conditions due to armed conflict, outbreaks of violence, and social/economic problems in affected areas complicated the public health response and increased the risk of disease spread both locally within the DRC and to other countries in the region such as Uganda, Rwanda, Burundi, Zambia, South Sudan, and Central African Republic (Riad et al., 2019; Schmidt-Sane et al., 2020; Wadoum et al., 2021).

On 20 September 2022, the Ministry of Health in Uganda, together with the World Health Organization - Regional Office for Africa confirmed an outbreak of EVD due to Sudan ebolavirus in Mubende District, Uganda, after one fatal case was confirmed. As of 19 October 2022, there have been 64 confirmed EVD cases, including 25 deaths (Case fatality rate: 39%; including probable deaths: 54%). Among these, at least 11 healthcare workers were infected, of whom five died. There are also 20 probable deaths among individuals who passed away before a sample could have been taken but who were linked to confirmed cases (European Centre for Disease Prevention and Control, 2022). This is the first Ebola disease outbreak caused by Sudan virus (SUDV) in Uganda since 2012 (World Health Organization, 2022b). The increased frequency of EVD outbreaks in DRC, Guinea and Uganda might be attributable to increased human-wildlife contact due to extensive deforestation, hunting, and mining practices (among other reasons), which calls for a robust monitoring system to inform future preventative policy measures.

Rwanda’s dense population and high-quality transport infrastructure make it highly susceptible to the rapid spread of EVD. Sharing borders with the DRC and Uganda increases Rwanda’s risk of cross border EVD transmissions due to the unmonitored movement of people and goods across the borders and weak border control systems. The increase in magnitude of the 2019 DRC EVD outbreak in Goma (in July 2019) and Mwenge Health Zone in South Kivu (in August 2019) further increased the risk of cross border EVD transmissions in Rwanda. Borders between Rwanda, DRC, and Uganda are particularly vulnerable as they are only separated by border check points. These borders, especially those shared with DRC, are characterised by intense trade activities and a high volume of population movement/migration. The threat of cross-border EVD transmissions is an emerging and increasing cause for concern not only for Rwanda but Burundi as well. Rwanda’s constant vulnerability to cross border EVD transmission necessitates strengthened EVD preventative and preparedness interventions, greater cross-border collaboration, and the flexibility to adapt to emerging needs.

The Government of Rwanda, through the Rwanda Biomedical Centre, launched the Multi-sectorial Risk Communication and Community Engagement Strategy for Ebola Virus Disease Preparedness and Response Strategy in August 2018. The strategy seeks to contribute to the national preparedness and response plan to halt EVD transmission in Rwanda, through effective evidence based social mobilization, community engagement and public education that advocates for social behavioural changes. Although Rwanda remains free of Ebola, this cost-effective control strategy requires an up-to-date understanding of peoples’ knowledge, beliefs, attitudes and behavioural patterns surrounding EVD. The overall goal of this survey was to understand the EVD-related knowledge, attitudes, and practices at individual, inter-personal, institutional, and societal levels.

## Methods

### Study Design

This cross-sectional survey was conducted to collect qualitative and quantitative data on the knowledge, attitudes, and practices surrounding EVD in five selected districts in Rwanda, i.e., Rusizi, Karongi, Rubavu, Burera and Gasabo districts (*Fig.1*). These five districts were purposely selected from the 15 districts identified as high-risk zones given their proximity (Rusizi, Karongi, Rubavu and Burera) or with air links (Gasabo) to DRC and Uganda.

**Figure. 1.**
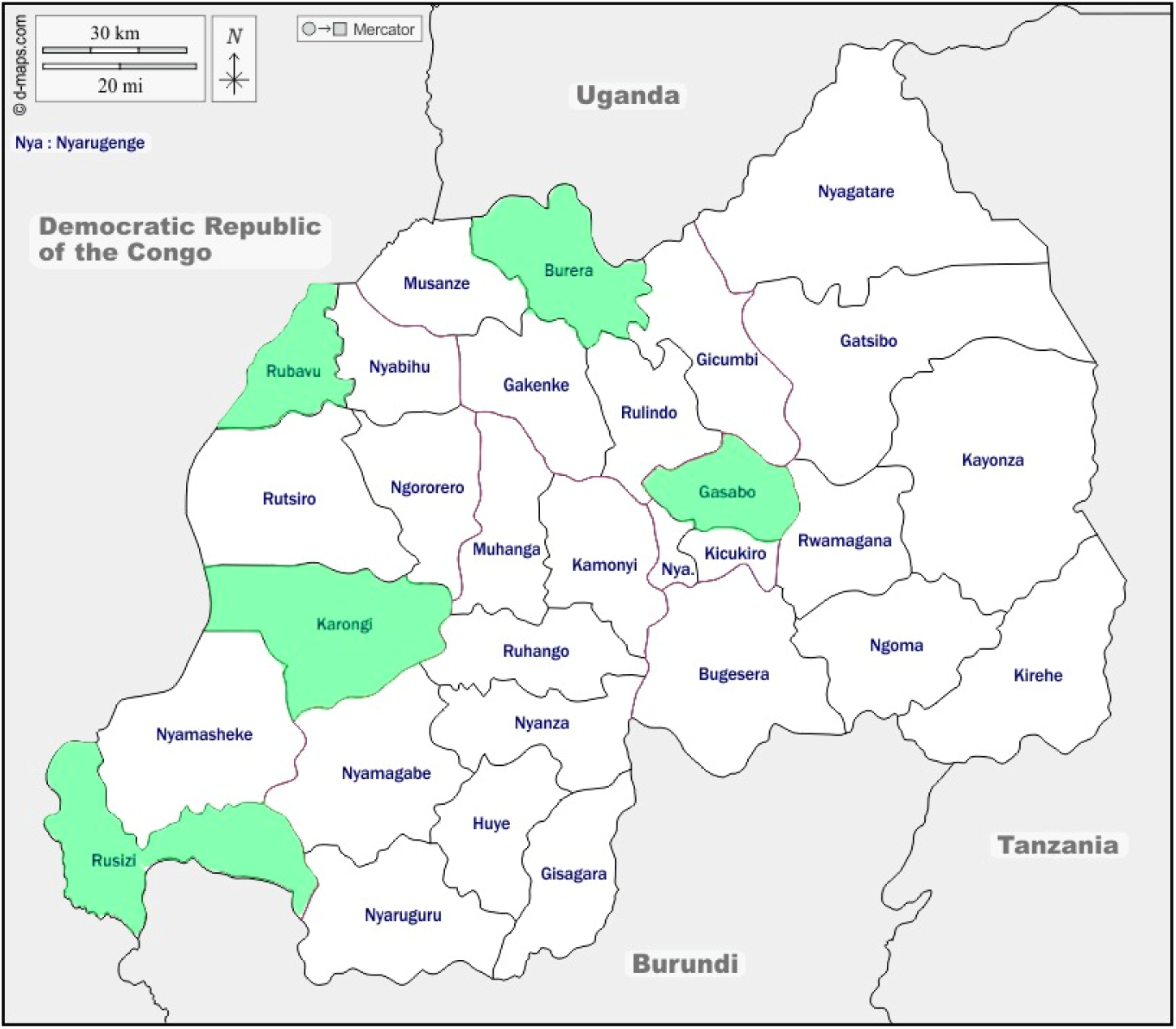
Map of Rwanda highlighting survey sites (green)

### Quantitative Approach

Due to the Government of Rwanda’s COVID-19 travel restrictions, all interviews were conducted over the telephone by a trained member of the research team. The research team consisted of two groups. The first group was composed of data collectors who were stationed in all five districts and sampled the participants, obtained informed consent, and collected participant contact details. The second group consisted of enumerators trained to administer the questionnaire. The enumerators called the participants and conducted the interviews over the telephone. Participants were randomly selected from urban and rural sectors in each village. The study participants were parents/caregivers or household members (male and female) of various backgrounds and occupations (including vendors, cross boarder traders, students, public transport drivers, security personnel, farmers etc.). Vulnerable groups such as child headed families, people living with disabilities, and others were deliberately selected and added to the study cohort. The sampling frame in each village was developed with the help of community leaders. Assuming 50% of the target population have an attribute of interest, a minimum sample of 1000 was found to be adequate at 95% confidence level, 4.95% level of precision and considering a non-response rate of 15%. In all five districts, the research team contacted a total of 1010 participants using a structured questionnaire: Gasabo (205), Karongi (203), Rubavu (207), Rusizi (193), and Burera (202). The questionnaire was translated and back-translated (English/Kinyarwanda) by two bilingual experts, and the questionnaire was modified according to their suggestions. The study team members validated the questionnaire. In order to assess the convenience and interpretation of the questionnaire, a pilot study was carried out on 35 participants from the general population and modified the questionnaire accordingly.

### Qualitative Approach

A purposive sampling strategy was used to select participants who participated in the Key Informant Interviews (KIIs). Participants from the Focus Group Discussion (FGD) were randomly selected from the list of contacts provided by the district-based data collectors. A total of 98 people participated through KIIs and FGDs. In each district, KIIs were completed with teachers, Early Childhood Development (ECD) care givers, Community Health Workers (CHWs) and Health workers in health centres. At the national level, KII participants were representatives from the United Nations Children’s Fund (UNICEF), Rwanda Biomedical Centre (RBC) and World Health Organization (WHO). All interviews were conducted over the telephone and Zoom platform. In each district FGDs were conducted with community leaders, parents/caregivers, and adolescents. Each FGD consisted of four people and were conducted through conference calls.

### Quality Assurance

The use of a conceptual framework and the development of data collection tools centred around the key research questions ensured data consistency and relevance. All of the tools incorporated questions developed around the conceptual framework to include knowledge, attitudes, and practices as well as the communication needs assessment of the target population. The research team also included public health experts, communication for development experts, and qualitative and quantitative researchers to ensure that all technical aspects of the study were satisfied. The team also included experts from the Ministry of Health who ensured that all processes conformed to national policies and protocols.

### Data Analysis

Quantitative data was collected using Kobo Collect (a real time data collection tool) and later transferred to the Statistical Package for Social Sciences (SPSS) v. 20 for analysis (Chicago, IL, USA). Findings of the survey were presented as proportions.

Qualitative data was managed using NVIVO 8 software. Interview transcripts were imported into NVIVO 8 for coding and subsequent analysis. To ensure inter-coder reliability and agreement, coding was performed by two researchers. Inter-coder agreement remained above 90% throughout the analysis.

### Ethical Clearance and Consent

Clearance to conduct the research was obtained from the Rwanda National Ethics committee (No.938/RNEC/2020) as well as the Rwanda National Health Research Committee. The research team received a training demonstration on obtaining informed consent and the principles of ethical research as well as guidelines to protect the research and study participants from COVID-19. Individual, written informed consent was obtained from participants and securely filed before joining the FGDs, KIIs, and household surveys.

## Results

### Quantitative findings

#### Characteristics of the study population

Out of 1010 respondents surveyed across five districts, 20.5% were in Rubavu, 20.3% in Gasabo, 20.01% in Karongi, 20.0% in Burera and 19% in Rusizi. In all surveyed districts, 56% of respondents were male, ranging from 49.2% in Rusizi to 60.4% in Rubavu. The highest proportion of respondents were married (70.3%), with 25.3% single, 3.1% widowed and 1.3% divorced. The majority of respondents reported that they had obtained a primary level of education (50%), followed by 33.7% with a secondary level of education, 7.5% University, 7.2% no prior education, and 1.6% tertiary level of education (Tab.1).

#### Assessment of knowledge and awareness on Ebola

Analysis of quantitative data suggests relatively high levels of awareness and knowledge pertaining to Ebola across the five study districts albeit with variations by district proximity to affected regions of DRC. Rubavu appears to have relatively high Ebola knowledge levels as the district is closest to the DRC, a country with the most reported Ebola cases. While the respondents in Gasabo indicated no official awareness regarding Ebola in their districts, participants in the district knew about Ebola primarily by means of radio communication, followed by TV and social media. Moreover, people seem to trust health professionals and community health workers as reliable sources of information. The overall Ebola knowledge levels disaggregated by district are shown in Table 2. The study participants responded that they receive Ebola-related awareness and knowledge from multiple sources, some of which (media sources) are prevalent across the population sub-groups.

**Table 1.** Characteristics of the study population, n=1,010

| ***Variable*** | ***Category*** | ***Gasabo*** | ***Karongi*** | ***Rubavu*** | ***Rusizi*** | ***Burera*** | ***Overall*** |
| --- | --- | --- | --- | --- | --- | --- | --- |
| ***n (%)*** |  | 20.3 | 20.1 | 20.5 | 19.1 | 20.0 |  |
| ***Sex*** | Male | 52.7 | 59.1 | 60.4 | 49.2 | 57.9 | 56 |
|  | Female | 47.3 | 40.9 | 39.6 | 50.8 | 42.1 | 44 |
| ***Education*** | No formal education | 14.1 | 6.4 | 8.7 | 4.1 | 2.5 | 7.2 |
|  | Primary | 50.2 | 49.3 | 36.7 | 56.5 | 57.9 | 50.0 |
|  | Secondary | 27.3 | 32.5 | 42 | 33.7 | 32.7 | 33.7 |
|  | Tertiary | 1.5 | 1.0 | 2.9 | 1.0 | 1.5 | 1.6 |
|  | University | 6.8 | 10.8 | 9.7 | 4.7 | 5.4 | 7.5 |
| ***Marital Status*** | Single | 29.3 | 11.8 | 29 | 32.6 | 24.3 | 25.3 |
|  | Married | 63.9 | 85.2 | 67.6 | 62.7 | 71.8 | 70.3 |
|  | Widowed | 3.4 | 3.0 | 1.4 | 4.1 | 3.5 | 3.1 |
|  | Divorced | 3.4 | 0 | 1.9 | 0.5 | 0.5 | 1.3 |

**Table 2.** Knowledge and awareness on Ebola

|  | *Gasabo* | *Karongi* | *Rubavu* | *Rusizi* | *Burera* | *Overall* |
| --- | --- | --- | --- | --- | --- | --- |
| ***Have you ever heard of Ebola*** |  |  |  |  |  |  |
| Yes | 100.0 | 99.5 | 100.0 | 98.4 | 100.0 | 99.6 |
| No | 0.0 | 0.5 | 0.0 | 1.6 | 0.0 | 0.4 |
| ***Perceived Causes of the Ebola Virus*** |  |  |  |  |  |  |
| Virus | 58.0 | 42.4 | 58.5 | 83.4 | 30.2 | 54.3 |
| Bats / monkeys / Chimps | 32.2 | 62.1 | 37.7 | 30.1 | 48.0 | 42.1 |
| Other | 6.4 | 15.3 | 2.4 | 2.0 | 15.8 | 8.5 |
| ***Transmission ways*** |  |  |  |  |  |  |
| Airborne* | 42.0 | 44.8 | 15.5 | 52.3 | 27.7 | 36.2 |
| Preparing bushmeat/Eating bush meat | 34.1 | 42.4 | 38.6 | 22.8 | 33.7 | 34.5 |
| Bodily fluids of infected persons | 72.2 | 53.7 | 79.2 | 48.7 | 51.0 | 61.2 |
| Mosquito Bite* | 7.3 | 1.0 | 2.4 | 2.1 | 4.0 | 3.4 |
| Shaking hands with infected persons | 40.5 | 39.9 | 74.4 | 30.1 | 45.5 | 48.3 |
| Using utensils used by infected persons | 5.4 | 36.0 | 32.4 | 25.9 | 22.3 | 24.4 |
| Other | 0.5 | 5.4 | 2.4 | 0.0 | 19.3 | 5.6 |
| Don't Know | 7.3 | 3.9 | 2.9 | 4.1 | 9.4 | 5.5 |
| ***Signs and Symptoms*** |  |  |  |  |  |  |
| Fever | 81.5 | 80.3 | 87.0 | 83.9 | 91.1 | 84.8 |
| Headache | 42.9 | 27.1 | 57.0 | 29.5 | 32.7 | 38.0 |
| Muscle pain | 19.0 | 10.3 | 30.4 | 9.8 | 20.8 | 18.2 |
| Diarrhoea | 34.1 | 43.3 | 47.3 | 33.7 | 33.7 | 38.5 |
| Abdominal pains | 15.6 | 3.0 | 25.6 | 9.8 | 11.9 | 13.3 |
| Sore Throat | 9.3 | 3.9 | 23.7 | 6.2 | 5.0 | 9.7 |
| Vomiting | 32.7 | 45.3 | 40.6 | 44.0 | 38.6 | 40.2 |
| Bleeding | 78.5 | 76.8 | 94.2 | 54.1 | 93.6 | 80.7 |
| Fatigue | 38.5 | 9.9 | 4.8 | 39.4 | 21.8 | 31.7 |
| Other | 3.9 | 1.9 | 1.9 | 1.0 | 5.9 | 3.5 |
| ***Prevention measures*** |  |  |  |  |  |  |
| Avoid contact with infected persons | 95.1 | 95.6 | 92.8 | 93.8 | 84.7 | 92.4 |
| Not eating uncooked bush meat | 36.1 | 41.9 | 34.8 | 19.2 | 29.7 | 32.5 |
| Bathing with salt and hot water* | 25.4 | 6.4 | 31.4 | 5.2 | 17.8 | 17.4 |
| By avoiding mosquito bites* | 4.4 | 0.0 | 10.1 | 4.1 | 3.0 | 4.4 |
| Not participating in burial rituals | 11.7 | 26.6 | 30.4 | 22.8 | 22.8 | 22.9 |
| *Not attending funerals* | 2.4 | 16.7 | 18.8 | 4.1 | 22.3 | 13.0 |
| *Other* | 4.4 | 10.8 | 10.6 | 1.0 | 30.2 | 11.5 |
| ***Received any training or health education on Ebola*** |  |  |  |  |  |  |
| *Yes* | 11.7 | 19.7 | 15.9 | 25.9 | 26.2 | 19.8 |
| *No* | 88.3 | 80.3 | 84.1 | 74.1 | 73.8 | 80.2 |
| ***Source of Information on Ebola*** |  |  |  |  |  |  |
| *Radio* | 95.1 | 93.1 | 92.2 | 96.8 | 92.6 | 93.9 |
| *TV* | 60.3 | 17.7 | 51.9 | 35.5 | 6.9 | 34.6 |
| *Social Media* | 23.0 | 16.3 | 29.1 | 17.7 | 6.9 | 18.7 |
| *Mobile Phone text messages* | 12.3 | 15.8 | 11.2 | 24.2 | 4.0 | 13.3 |
| *Health workers* | 18.6 | 23.2 | 18.4 | 31.7 | 23.3 | 22.9 |
| *Flyers/Posters* | 12.7 | 2.0 | 17.5 | 16.7 | 11.9 | 12.1 |
| *Church/Religious workers* | 6.4 | 17.2 | 5.8 | 7.0 | 10.4 | 9.4 |
| *Traditional healers* | 0.5 | 0.0 | 1.0 | 0.0 | 0.0 | 0.3 |
| *Email* | 0.0 | 1.0 | 1.5 | 2.7 | 0.0 | 1.0 |
| *Drama* | 4.4 | 2.0 | 0.0 | 0.0 | 0.5 | 1.4 |
| *Newspapers* | 11.3 | 1.0 | 4.4 | 16.7 | 4.0 | 7.3 |
| *CHW* | 9.8 | 23.6 | 20.4 | 22.0 | 26.2 | 20.4 |
| *Other* | 1.0 | 2.5 | 4.9 | 1.1 | 21.8 | 6.3 |
| ***Trusted Sources of Information*** |  |  |  |  |  |  |
| *Health professionals* | 71.1 | 56.7 | 68.4 | 69.1 | 60.4 | 65.1 |
| *Community health workers* | 23.0 | 33.5 | 28.6 | 22.3 | 26.2 | 26.8 |
| *NGO* | 2.5 | 0.5 | 0.0 | 4.3 | 0.0 | 1.4 |
| *Traditional leaders* | 0.0 | 0.0 | 0.0 | 0.5 | 0.0 | 0.1 |
| *Religious leader* | 0.0 | 1.0 | 0.0 | 1.6 | 0.5 | 0.6 |
| *Other* | 3.4 | 8.4 | 2.9 | 2.1 | 12.9 | 6.0 |

\*Wrong response

**Table 3.**
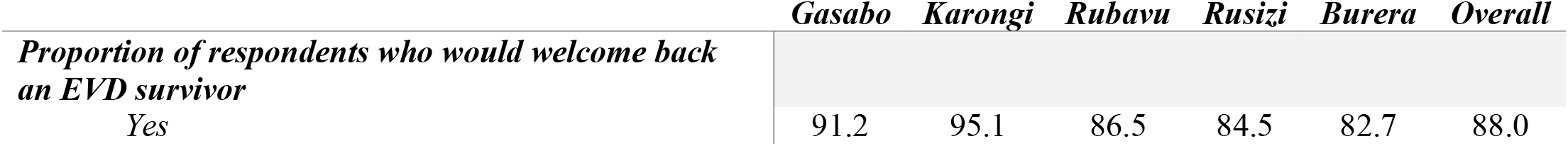
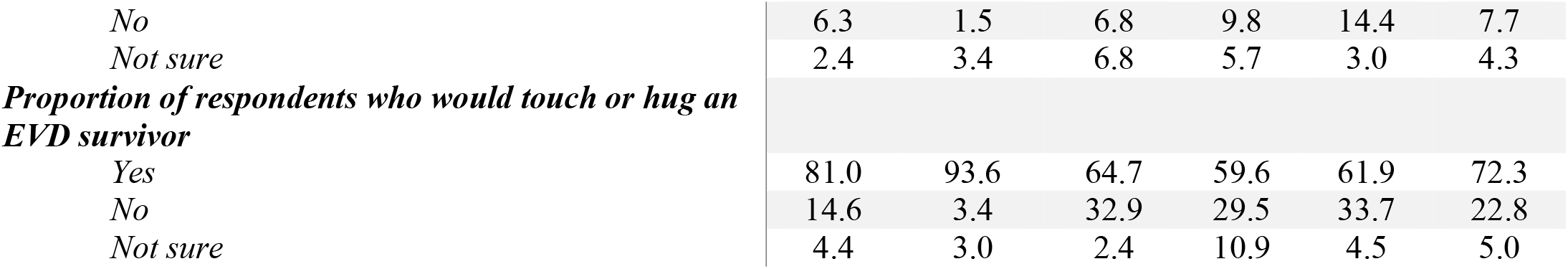
Knowledge attitudes towards Ebola infected and affected persons

#### Assessment of attitudes towards Ebola infected and affected persons

Overall, the respondents in all districts showed a positive attitude towards EVD survivors. For instance, 88% of surveyed respondents reported that they would welcome back a neighbor who survived Ebola. The percentage of respondents indicating this positive attitude was highest in Karongi (95.1%) and lowest in Burera (82.7%). The table below shows the proportion of respondents and their responses regarding welcoming back into their community, touching or hugging an EVD survivor.

#### Assessing the risk perceptions and beliefs about Ebola

The quantitative data analysis indicated a heightened level of Ebola risk perception in Rwanda as a country because it neighbours DRC, which is the EVD epicentre in the East African region: An average 90% of all the respondents believe that Rwanda was at risk of an Ebola outbreak and 87.8% of the participants think that, individually, they are at risk. Furthermore, most of the respondents do not seem to believe in traditional/spiritual-based therapies for treating EBV (91.5% and 89.6%, respectively. Table 4 shows the respondent risk perceptions and beliefs about Ebola.

**Table 4.**
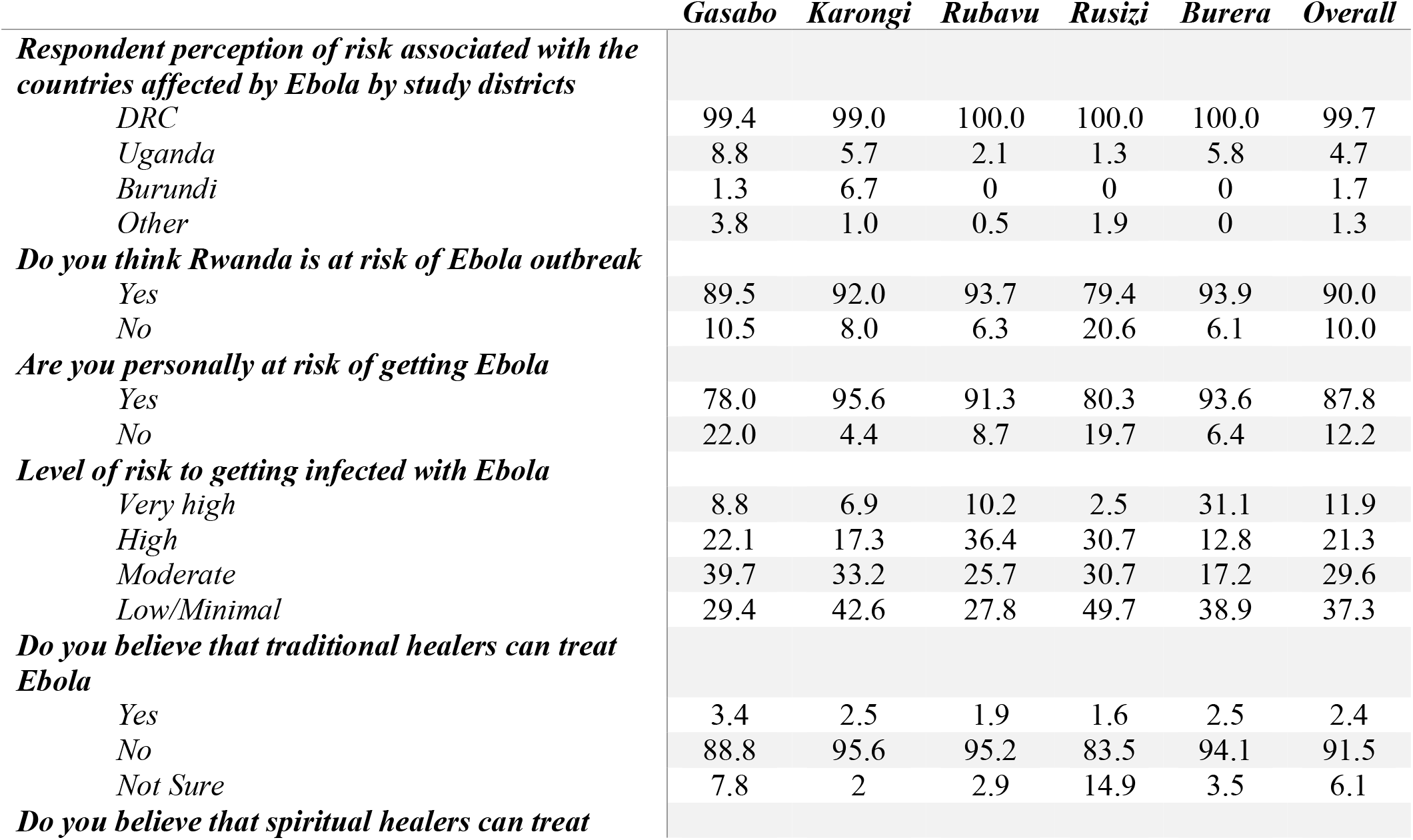
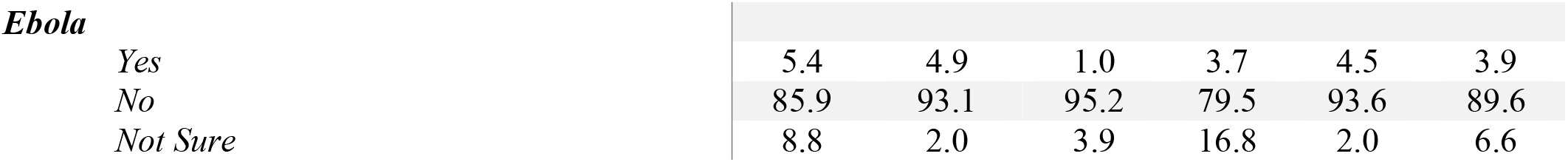
Knowledge the risk perceptions and beliefs about Ebola

#### Assessment of Ebola related behaviours and practices

The respondents were also asked to indicate the action they would take for themselves and family members to avoid Ebola. Table 5 presents the proportion of respondents with respect to various personal hygiene practices to avoid Ebola. The most common preventive measures which were mentioned in each of the locations were hand washing and limiting body contacts. The results also indicated that 97.2% of respondents believed that it is important to get vaccinated against Ebola virus. In fact, 73.6% answered “yes” for changing their behaviour or habits to avoid infection.

**Table 5.** Knowledge of Ebola related behaviours and practices

|  | *Gasabo* | *Karongi* | *Rubavu* | *Rusizi* | *Burera* | *Overall* |
| --- | --- | --- | --- | --- | --- | --- |
| ***Personal Behaviours / hygiene practices to avoid Ebola.*** |  |  |  |  |  |  |
| *Practice handwashing and good personal hygiene* | 86.8 | 87.7 | 91.8 | 83.4 | 85.6 | 87.1 |
| *Avoiding touching sick people or the dead* | 75.1 | 65.0 | 65.7 | 81.3 | 54.5 | 68.2 |
| *Not eating raw bush meat* | 15.1 | 28.6 | 15.9 | 17.6 | 23.8 | 20.2 |
| *Washing with salt and hot water* | 20.5 | 0.0 | 14.5 | 2.6 | 5.4 | 8.7 |
| *Other* | 2.9 | 7.9 | 8.2 | 0.5 | 19.3 | 7.8 |
| ***Have you changed your behaviour or practices to avoid being infected with Ebola?*** |  |  |  |  |  |  |
| *Yes* | 60.5 | 79.8 | 91.8 | 69.9 | 65.3 | 73.6 |
| *No* | 34.6 | 4.4 | 6.8 | 15.0 | 33.7 | 18.9 |
| *Not sure* | 4.9 | 15.8 | 1.4 | 15.0 | 1.0 | 7.5 |
| ***Behaviours in the event a family member contracted Ebola*** |  |  |  |  |  |  |
| *Do nothing* | 0.5 | 0.0 | 0.0 | 0.0 | 0.0 | 0.1 |
| *Hide the sick person* | 1.0 | 0.0 | 0.0 | 0.0 | 0.0 | 0.2 |
| *Isolate the sick person* | 13.7 | 3.0 | 21.3 | 5.3 | 5.9 | 9.9 |
| *Call the hotline* | 53.7 | 68.0 | 54.1 | 28.6 | 61.4 | 53.5 |
| *Take patient to health centre or hospital* | 21.0 | 16.3 | 18.8 | 38.6 | 13.9 | 21.5 |
| *Take patient to an Ebola Treatment Centre* | 9.8 | 2.5 | 3.9 | 27.5 | 2.0 | 8.8 |
| *Other* | 0.5 | 10.2 | 1.9 | 0.0 | 16.8 | 6.0 |
| ***Do you believe that it is important to get vaccinated to prevent Ebola*** |  |  |  |  |  |  |
| *Yes* | 97.6 | 96.6 | 99.0 | 95.3 | 97.5 | 97.2 |
| *No* | 2.4 | 3.4 | 1.0 | 4.7 | 2.5 | 2.8 |
| *Do you believe that vaccines against Ebola are safe* |  |  |  |  |  |  |
| Yes | 97.1 | 92.1 | 87.0 | 86.0 | 88.1 | 90.1 |
| No | 2.9 | 7.9 | 13.0 | 14.0 | 11.9 | 9.9 |

### Qualitative findings

#### Assessment of knowledge and awareness on Ebola

Analysis of qualitative data suggests there are variations on overall knowledge on Ebola among sub-population groups (Health Workers, Community Leaders, Adolescents and Teachers for both ECD and Secondary schools).

> *We have learned that Ebola is caused by the virus which has its origin in animals:* **FGD, Community Leader Rubavu (Rural)**.
>
> *I do not know the causes of Ebola, but I know its symptoms like vomiting, bleeding and strong fever, things like that*. **KII, Teacher Karongi (Urban)**.
>
> *I wish I had knowledge on Ebola. I could explain to students, I do not have enough knowledge. I am not ignorant completely. I know little about it such as what we hear from radios on the nature of the epidemic*. **Teacher Karongi (Rural)**.

Although Rwanda has not yet recorded an EVD outbreak, the study respondents were knowledgeable regarding the prevention of future outbreaks, and the knowledge was almost uniform across the districts, study population sub-groups, and geographical areas.

> *Yes, Ebola is preventable. To me first thing is cleanliness, if we know it is spread through liquids, we can avoid touching one another and this can help*. **KII, Teacher, Karongi (Rural)**.
>
> *Prevention of Ebola is washing hands regularly, avoiding crowded places or places where there is an outbreak of Ebola*. **FGD, Parent Caregiver, Burera (Rural)**.
>
> *We can prevent Ebola through prevention measure to include washing hands, avoiding assembling, avoiding shaking hands and more important getting vaccinated against Ebola*. **FGD, Adolescent, Rubavu (Urban)**.

Overall the respondents reported receiving knowledge on Ebola mainly from their community leaders and community health workers.

> *During community work called “umuganda” health workers are invited to share information on Ebola. However, since March 2020, there has not been any such educative talks on Ebola*. **FGD, Parent/Caregiver, Rusizi (Rural)**.
>
> *In Rusizi, there is a programme of vaccination against Ebola, so community leaders have information on Ebola. Also doctors from Kigali help people in the district on how to take measures to prevent Ebola*. **FGD, Community Leader, Rusizi (Urban)**.

Several districts have been organizing the awareness campaigns to educate the public about how to better prevent the spread of the Ebola disease. For instance, community leaders in Rubavu indicated that they had received formal Ebola training from health workers at the district level as a part of the district awareness raising strategies.

> *All Community leaders and opinion leaders have been trained and involved in Ebola education. The training was an alert in Rubavu district. We even did campaigns and the cars were driving around the town with loudspeakers with instructions on knowledge on Ebola and preventive measures*. **FGD, Community Leader, Rubavu (Urban)**.

The analysis of the feedback from participants identifying as teachers across the study districts noted little to no formal (or informal) Ebola-related dissemination of knowledge and/or training for teachers. For instance, teachers in Karongi reported limited exposure to educative activities on Ebola-related knowledge. One of the teachers had this to say:

*Honestly, I do not know much about Ebola. We simply just get some information from the radio, but this is not comprehensive for the role we play as teachers who should impart such knowledge to the children, who would in turn also share the information with their families at home*. **KII, Teacher, Karongi (Urban)**.

#### Assessment of attitudes towards Ebola infected and affected persons

Responses from the FGDs with parents and local leaders indicated that in general, the community members have a positive attitude towards survivors. The following is an excerpt from one of the participants.

> *Sometimes the person can be quarantined. However, when he or she is sent to the hospital and the hospital discharges him/her because he/she is cured, then there is no reason to fear or quarantine him/her. We can welcome him/her back*. **FGD, Parent/Caregiver, Karongi (Urban)**.

Conversely, the results also suggest that a significant proportion of respondents have negative attitudes towards Ebola survivors should it affect the country, as a whole. These results are also supported by information gathered through FGDs. Parents and local leaders who participated in FGDs in all five districts also indicated that they could eat, hug, and shake hands with a person who has recovered from Ebola. However, some local leaders revealed that some people need more time to see if everything is fine with the survivor.

> *For the recovered member, it takes time to go back into the community, the population would like to be sure that he has fully recovered, but there is no discrimination*. **FGD, Local Leaders, Rubavu, (Rural)**.

Parents and local leaders also reported a willingness to adhere to Ebola burial guidelines and protocols when burying someone they suspect that he has died of Ebola (i.e., no touching the corpse and allowing the burial team to come and bury body). The following are extracts from the FGDs conducted with the parents.

> *For someone who died of Ebola, there is a burial team and because of fear of Ebola no one can approach the dead body, the community accept use of burial team, even if families are psychologically affected by not paying the last respects to the dead*. **FGD, Parents/Caregivers, Gasabo (Urban)**.
>
> *In our customs we are used to make the last respect to the dead person, so it is too hard to accept the burial team, but it is a must, sometimes one member of the family can wear the protective equipment and be part of the burial team*. **FGD, Local leaders, Rubavu (Rural)**.

A large proportion of the population in all districts is still afraid or apprehensive of survivors. National level KII noted the need for more sensitization and recommended that there should be a refresher conducted on EVD to raise awareness.

> *My immediate suggestion would be to conduct a refresher related to Ebola especially when it comes to community sensitization approaches, latest information, latest science about Ebola, strengthen the knowledge of community health workers about the vaccine. Also because of turnover, this refresher will be new information for new health workers. We should also have a branded corner in health facilities where people can access Ebola information. We also need to engage teachers, ECD caretakers, young children to institutionalize knowledge about Ebola, plus we should constantly update to highlight the difference between Covid and Ebola. We also need to have a refresher for religious leaders*. **KII, UNICEF, Gasabo (National)**

#### Assessing the risk perceptions and beliefs about Ebola

Most of the respondents think that Rwanda was at risk of an Ebola outbreak. This is reflected in the FDG/KII findings with respondents citing both DRC and Uganda as the source of regional Ebola cases. Respondents also believe cross boarder-related activities including trade, free movement, and fishing (especially on Lake Kivu) increase Rwanda’s risk to Ebola.

> *“Yes, the epidemic can come in Rwanda because our neighbour DRC is regularly suffering from the epidemic and we are at a risk since there can happen to a person from DRC who can for instance enter and contaminate or spread the Ebola”*, **KII, Teacher, Gasabo, (Urban)**.
>
> *Many people in the community know the seriousness of Ebola. Before Covid-19, hospitals sent Health workers into different churches to teach them how to take measures against Ebola virus”*. **FGD, Community Leaders, Karongi, (Rural)**.

The fact that neighbouring countries such as DRC and Uganda had reported Ebola cases put the Rwandans at high risk. The following excerpts suggest the potential risk of Ebola as a result of illegal movement between boarders.

> *Where we live, there are people who come from DRC to pick sand. We need to avoid contact with these people. They are picking sand at a place called Kiraro*. **FGD, Adolescent, Karongi (Urban)**.
>
> *With new DRC, one method of transport is boat. Many people come from Goma and Bukavu and there it is possible for Rwandans to get Ebola. Also, here in Karongi, fishing is an important business. The lake is between two countries, but boarders are not clear and there is constant movement between these people doing this business and other businesses which make the spread of Ebola possible*. **FGD, Community Leader, Karongi (Urban)**.
>
> “*It has been said Ebola is coming from bats and in Rwanda we have those bats. So, it can come also. In addition, movement from Goma to Rwanda can facilitate the spread of the virus*”. **FGD Adolescents, Gasabo (Urban)**

The majority of the respondents from the FGDs and KIIs perceive that everyone in Rwanda takes Ebola as a very serious disease that spreads. However, isolated responses from FGDs with adolescents in Burera, and parents in Gasabo, Karongi, Rubavu and Burera mentioned that Ebola was a political disease, one that is ‘made-up’ by politicians.

> *“Many people in the community know the seriousness of Ebola. Before Covid-19, hospitals sent Health workers into different churches to teach them how to take measures against Ebola virus”*. **FGD, Community Leaders, Karongi (Rural)**
>
> *“Once I heard traders talking about a political disease”*, **FGD, Adolescent, Burera (Rural)**.
>
> *“People have different way of understanding. Some people have the misconceptions on Ebola. Some said that Ebola is a political disease, it does not exist”*. **Parent FGD, Gasabo (Urban)**.

The majority of myths and misconceptions were cited in KIIs and FGDs conducted in Gasabo and the majority were captured in the KIIs with health workers. The persistence of myths and misconceptions also indicates a need for EVD awareness campaigns.

> *“The myths and misconception about EVID, it is said that the epidemic is caused by a virus made from laboratories and brought in Africa”*. **KII, Health Worker, Gasabo (Urban)**.
>
> “*It is somehow said that it is an antichristian epidemic and aiming at imparting satanic stamp of 666*”. **KII, Health Worker, Gasabo (Urban)**.

#### Assessment of Ebola related behaviours and practices

Respondents were asked to indicate whether they had changed their behaviour or practise to avoid being infected with Ebola. This change in behaviour was attributed to the community meetings/umuganda that were held before COVID-19 restrictions. At these meetings the members of the community were trained and equipped with various methods to mitigate contracting the disease. These training sessions were performed by health workers who gave speeches and handed out materials at these events.

> *During the community work also, we were given the opportunity to talk to the people. When you suspect a person, you mention it on his transfer then you wait for the answer from the health centre to see if you were right*. **KII, Health worker, Karongi (Urban)**

In the event of onset of symptoms or confirmation of an Ebola case within a family, respondents across all the groups (FGDs & KIIs) cited isolation as the recommended practice. Interestingly, parents in Gasabo went on to mention that isolation should be followed with testing of all family members in case of confirmation of an Ebola case.

> “*In my opinion, in case families suspect Ebola, we have to inform our community health workers in my village, because I know, they have more information on health issues. Usually, we always seek help from community health workers because they are close to us and know how to behave in case of illness, we trust them, but in the meantime, we have to isolate the suspect*”. **FGD, Parent/caregiver, Gasabo (Urban)**.
>
> “*When we suspect one of member to be infected better to call the health workers while avoiding any contact and approach to that person*”. **FGD, Parent/caregiver Rusizi (Rural)**.

Some respondents indicated that they were previously vaccinated or willing to be vaccinated, highlighting the importance of EVD vaccines in preventing further infections and death. The vaccinated participants attested to experiencing no side-effects from taking the EVD vaccine.

> *I saw many being injected especially those bordering Democratic Republic of Congo and I was wondering when we will get the same vaccines too, unfortunately we did not qualify for the first round, but many are ready for the EVD vaccines*. **FGD, Community Leader, Karongi (Rural)**.
>
> *I have already been vaccinated and my family. If there are preventive measures, they are not 100 percent tight. The only way to be definite is to get vaccinated*, **FGD, Community Leader, Rubavu (Urban)**.
>
> *My family has been vaccinated for Ebola. We did not experience any problem. We think the vaccine is safe because it is tested on those that received it*. **FGD, Community Leader, Rubavu (Urban)**.

Most of the respondents were with the opinion that they saw no reason for communities to refuse vaccination. A few participants, however, cited myths and rumours (microchip or causing infertility) as the cause for hesitancy but also believed this could be overcome through awareness campaigns and community mobilisation.

> *Nothing in my community can prevent people to get vaccinated, sometimes rumours but with sensitization it was over* **FGD, Community leader, Rusizi (Rural)**.

The channel of communication that is most preferred by the respondents is radio, and in some cases Television (TV), but radio stands out across key informants and focus groups.

> *“The Radio is the principal means since almost everyone has it. You can invite people in the community for a meeting and few will come but when it comes to the announcement made on the radio, everyone will attend*.*”* **KII, Teacher, Karongi (Urban)**
>
> *“Primarily they (communities) listen to the radio, they watch television and also listen to cars going around with big speakers*.*”* **KII, Health Worker Rubavu (Rural)**

When it comes to trusted sources of information, the respondents collectively viewed the Ministry of Health and its personnel, Government Institutions, such as Rwanda Biomedical Centre, and local leaders as being more trustworthy in times of disease outbreaks.

> *“Those who do meetings like Country leaders at the country level. We cannot accept the information from the district unless we hear ourselves from high rank officials. Unless it comes from the cabinet, otherwise we can’t trust the rest*.*”* Teacher, Karongi (Rural)

Some of the challenges regarding Ebola communication are connected to the current focus on the COVID-19 pandemic. COVID-19-related communication and messaging is the number one public health priority, as well as the effects of COVID-19 related restrictions on mass gatherings that have rendered interpersonal and group communications virtually impossible. One health worker had this to say:

> *“the challenge is that we no longer have public gatherings, therefore it is not easy to reach a health worker for some. Sometimes the village (is) also big, which means it can take almost a month to cover it*.*”* **KII, Health worker Karongi (Urban)**

Other challenges that have been raised are rumors and inaccurate dissemination of information, and the long distances that individuals in rural settings have to travel to seek appropriate medical attention. Information from national level Key informants indicated that there is need to strengthen the flow of educational information about Ebola. One of the key informants said the following.

> *What is important is to reinforce the flow of information so that there is no cut in the communication to ensure that the right information is in the right place at the right moment. we monitor this very closely also to make sure that there are no rumours*. **KII, WHO, Gasabo (National)**.

## Discussion

This study sought to describe the knowledge, attitudes and preventive practices concerning EVD in five selected districts at risk of cross-border spread, four bordering DRC or Uganda and one with air links to DRC and Uganda. Apart from few unpublished technical reports from the Ministry of Health, this is, to the best of our knowledge, the first comprehensive analytical KAP survey on EVD in Rwanda. Such a study is essential for developing preventive interventions for community members considering the increased risk of cross border EVD spread between DRC, Uganda and Rwanda. The risk of cross border EVD spread is exacerbated by the high volume of people traveling across the Rwanda-DRC and Rwanda-Uganda borders.

We observed relatively high Ebola-related knowledge and awareness levels across the five study districts albeit with variations by district proximity to affected regions of DRC and by population subgroups. Over 99% of respondents in the five study districts had heard of Ebola. Such a high level of awareness is a good indication of the awareness raising efforts that have been previously undertaken by members of the Government of Rwanda through its Ministry of Health. Consultations with health workers across the five districts indicated that they had received training on Ebola prior to the COVID-19 pandemic, which had covered topics on general knowledge surrounding Ebola, such as what the causative agent is, how it is spread, how it can be prevented, as well as the hallmark signs and symptoms. The training is likely to have contributed to a relatively high level of knowledge on Ebola. Apart from this training for health workers, most respondents across the five districts reported that they mainly received the information on Ebola through media sources, including radio and TV. Another proportion of respondents reported receiving information on Ebola from posters/notice boards at health facilities. People who came from DRC were also a source of knowledge for the adolescents. The channel of communication that is most preferred among respondents from all districts is radio, which may be attributed to its increased accessibility (when compared to TV or social media). This finding, while not surprising, underscores the importance of radio in the context of health-related communication, especially in rural areas where other media sources are less accessible.

Despite the generally high level of Ebola-related knowledge, gaps remain in respondents’ knowledge and attitudes towards EVD that were likely to impact preventive practices and behaviours. This was observed in survey results where a large proportion of people (as high as 23%) were apprehensive about whether they should physically and/or socially interact with survivors. For example, about 22.8% were not sure whether they would hug or touch a survivor or not. A small percentage indicated that they would not have a personal or close interaction (eat, work, study together, hug or touch) with EVD survivors. These discriminatory tendencies are the result of the EDV myths and misconceptions that are being perpetuated in most analysed study areas and particularly in rural areas. The most common misconceptions leading to the discrimination of survivors relates to how one can be infected with EVD, and the origin of the virus. EVD-related discrimination is largely based on community fear that EVD survivors are still contagious (James et al., 2020). Such discrimination has led to EVD survivors being mocked by their communities, being evicted from their homes by their property owners, losing their former jobs, and being divorced by their spouses (James et al., 2020; Karafillakis et al., 2016; Rabelo et al., 2016; Schwerdtle et al., 2017; Sow et al., 2016). Most of our study participants indicated that one’s re-acceptance into the community following EVD infection depends largely on being able to provide a certification of treatment and recovery from an Ebola treatment centre. Previous studies have shown that some EVD survivors were restricted from visiting public places, such as public toilets, and have experienced difficulty in trading commodities at their local market due to a community reluctance to touch their items or money (James et al., 2020; Rabelo et al., 2016). These results demonstrate that comprehensive advocacy and awareness programs are still needed. Limited knowledge about Ebola treatment could also be one of the biggest drivers of the negative attitudes channeled towards individuals who previously recovered from Ebola (Carter et al., 2017; Pham et al., 2022).

Ebola virus disease was historically perceived as a near certain killer in the absence of treatment; however, that is no longer the case. Supportive care, rehydration with oral or intravenous fluids, and treatment of specific symptoms are now used to improve the likelihood of survival. In addition, two monoclonal antibodies (Inmazeb, REGN-EB3 and Ebanga, MAb114) were approved for the treatment of *Zaire ebolavirus* (Ebolavirus) infection in adults and children by the US Food and Drug Administration in late 2020 (CDC, 2019; World Health Organization, 2021b). The provision of supportive medical care for EVD infected patients, combined with monoclonal antibody treatment now lead to recovery for the vast majority of individuals. For full community-level protection, gaps in Ebola-related knowledge and unrestricted access to treatment must be overcome, especially in resource-poor areas.

The findings of this study indicate that various groups of people are at high risk of contracting and spreading Ebola in Rwanda. The categories include immigration officers, customs staff, cross border traders, and health workers. The health personnel are at increased risk, especially as EVD infected individuals present with symptoms consistent with other known diseases such as malaria and the appropriate levels of caution and prevention are not taken. In combination, these high risk groups in EVD-affected areas should take special care in minimizing their risk of exposure.

Variability in behavioural practices was observed by district. For instance, residing in closer proximity to affected areas resulted in residents being more alert in ensuring they protected their family members from contracting the virus. The most common communal practices are centred around the sharing of information about Ebola prevention. This may be by forming clubs to educate others on Ebola prevention practices, contacting relevant health authorities through an emergency line (i.e., 114) and reporting suspected cases to the nearest health facility. Due to the high CFR, EVD undoubtedly instils fear in the inhabitants of any affected community. According to previous studies, EVD-related fear influences inhabitants to act with increased caution and to resort to preventive measures against the disease. These changes in human behaviour resulting from fear of the virus may lead to a decrease in the human to human, and human to pathogen contact rates (Juga et al., 2021).

About 90.1% of respondents consulted during the study felt that increased personal protection could be achieved via vaccination and that the EVD vaccine was safe. The Government of Rwanda, in collaboration with WHO and other partners, have been implementing public health measures to protect its population against Ebola Virus disease. Vaccination was one of the measures taken to protect health care and frontline workers in the districts at high risk of Ebola. Overall, 2874 HCWs and Frontline Workers from the highest risk districts were vaccinated in 2019. Additionally, Rwandan residents residing near the border with the DRC were offered the Ad26.ZEBOV (adenovirus type 26 vector vaccine encoding Ebola virus glycoprotein) and MVA-BN-Filo (modified vaccinia virus Ankara vector-based vaccine, encoding glycoproteins from Ebola virus, Sudan virus, and Marburg virus, and nucleoprotein from Tai Forest virus) Ebola vaccine regimen (Nyombayire et al., 2022).

## Conclusion

Overall, the study noted relatively high knowledge levels among the study respondents with 99.6% reporting having heard of Ebola. Such high level of awareness is a strong indication of the awareness raising efforts that have been previously undertaken by the Government of Rwanda. The results of this study also suggest that future communication and media efforts should be focused on knowledge dissemination in the public domain, such as available treatment options, case reporting, and positively interacting with EVD survivors. These findings, along with existing data and previous experience in communicating Ebola throughout the country should help guide other high-risk countries in creating an effective, evidence-based framework for controlling EVD.

### Limitations and Mitigation Strategy

The outbreak of the COVID-19 pandemic and the Government of Rwanda’s COVID-19 restrictions forced changing data collection methods from face-to-face interviews to virtual interviews. Although contextually expedient, conducting interviews virtually created unforeseen challenges, where roughly 50% of selected respondents refused to provide informed consent. Consequently, the research team was required to recruit more participants to replace those who had chosen not to participate in the study.

## Data Availability

For ethical reasons raw data generated during the current study were not made publicly available. Access to the datasets analysed may be facilitated upon reasonable request to the corresponding author.

## Acknowledgments

We thank all the study respondents for their generous time and patience, and for voluntarily sharing their personal experiences with us. We would also like to express our appreciation to Mr Maksim Fazlitdinov, Mr Fulgence Kamali, Mrs Balisanga Helen, Mrs Perpetua Kububa, Mr Valens Nkurikuyinka, Mr. Chikondi Khangamwa, Mr Kenneth Mulondo, Mr Leonard Mbwanda, Mr Innocent Vomitadyo, Dr Nedy Matshalaga, Eng. Simukai Sibanda for their assistance in the implementation of this study.

This work was supported by grants from United Nations Children’s Fund (UNICEF) Rwanda, Canadian Institutes for Health Research, Genome Canada/Atlantic Genome, Research Nova Scotia, Dalhousie Medical Research Foundation, and the Li-Ka Shing Foundation. A technical working group composed of the University-based Researchers, UNICEF representatives, and the Rwanda Ministry of Health provided input from the inception through to publication of this manuscript. Nikki Kelvin (The Journal of Infection in Developing Countries) and Jean Claude Semuto Ngabonziza (Rwanda Biomedical Center, Kigali, Rwanda) provided editing assistance throughout the preparation of this manuscript.

